# Analyses and Forecast for COVID-19 epidemic in India

**DOI:** 10.1101/2020.06.26.20141077

**Authors:** Rudra Banerjee, Srijit Bhattacharjee, Pritish Kumar Varadwaj

## Abstract

COVID-19 is a highly infectious disease that is causing havoc to the entire world due to the newly discovered coronavirus SARS-CoV-2. In this study, the dynamics of COVID-19 for India and a few selected states with different demographic structures have been analyzed using a SEIRD epidemiological model. A systematic estimation of the basic reproductive ratio *R*_0_ is made for India and for each of the selected states. The study has analysed and predicted the dynamics of the temporal progression of the disease in Indian and the selected eight states: Andhra Pradesh, Chhattisgarh, Delhi, Gujarat, Madhya Pradesh, Maharashtra, Tamil Nadu, and Uttar Pradesh. For India, the most optimistic scenario with respect to duration of the epidemic shows, the peak of infection will appear before mid September with the estimated *R*_0_ = 1.917, from the SEIRD model. Further, we show, a Gaussian fit of the daily incidences also indicates the peak will appear around middle of August this year. Our analyses suggest, the earliest dates when the epidemic will start to decline in most states are between Jun-August. For India, the number of infected people at the time of peak will be around 1.6 million including asymptomatic people. If the community transmission is prohibited, then the epidemic will infect not more than 3.1 million people in India. We also compared India’s position in containing the disease with two countries with higher and lower number of infections than India and show the early imposition of lockdown has reduced the number of infected cases significantly.

## 1. Introduction

COVID-19 was announced as a global pandemic by World Health Organization (WHO)[1] owing to its highly contagious and pathogenicity that has been rapidly spreading throughout the world since its first reported outbreak in China in December, 2019. This highly contagious virus is spreading in a much higher rate than the other earlier reported corona viruses. In India, the first case of COVID-19 was reported on 30 January, 2020 and even after 23 weeks of its existence and four phases of lock-down, the daily rise of cases is alarming. To manage an epidemic like this, the administrations and health departments across the country like India with 1.35 billion population, need a more or less clear idea of the challenges they need to tackle during the course of epidemic. Also a forecasting about the said outbreak can help in decisive planning and management of the resource allocation. Therefore a detailed analysis based on real data should prove to be very much useful to estimate the possible amount of testing kits, hospitalizations, quarantine centres etc. needed to contain the disease.

The motivation behind this work is to forecast the situation with the help of SEIRD population model, a well established model to study the dynamics of infectious diseases. This may be useful for the health officials and administration to estimate how they should be prepared for coming days to provide treatment and containment measures for COVID-19. The assessment for different states will also show the trend of the epidemic. With the sufficient amount of data in our hands, we can nowcast and forecast the possible situations that a state or the whole country is going to face. India invoked complete nationwide lockdown in the early phase of the epidemic on March 25th. This has prevented rapid transmission of the virus among people and also has given time to the administration and health officials to prepare for the measures that needed to be taken to tackle the situation. However, an early lockdown for a country with over 13 billion people has slowed down the pace of the spread of the virus considerably and resulted in a delayed appearance of flattening of the temporal progression of the infected people. It is thus important to find how long should the countrymen remain alert and careful about the spread of the virus.

For prediction of the progression of epidemic, we need a well structured data. The accurate estimation of model parameters would give us most plausible situation. Although the data published by ICMR, and other agencies are quite well organised but not sufficient to compute certain parameters accurately. In this situation we have two options. Either we take help from estimates of parameters from other countries where a well organised data was used or we can take data of various states in India with different demographics, compute the parameters for each of them, and compute the values of those parameters to find an estimate the parameters. We have chosen 3 states with high population density (Delhi, Tamilnadu, Uttar Pradesh), three states with medium population density (Andhra Pradesh, Maharashrtra, Gujarat) and two states of lower population density (Madhya Pradesh, Chhatisgarh). These states are also further divided into high, moderate and low infection rates. We have employed same methodology for each of these states and projected the values of the different parameters and executed the Susceptible-Exposed-Infected-Recovered-Dead (SEIRD) model to see the trend of COVID-19 transmission. The trends are then compared with the existing data for those states, which in turn enhance predictability.

The recently reported work [2] has compared the whole genome sequence of 28 viral strains from India. The analysis has reported a novel non-synonymous mutation in NSP3 gene of SARS-CoV-2 along with other frequent and important mutations reported across the globe. This suggests the possibility of having different strains at different part of India looking into the demographic density, travel history, inflow of migrant labours, cultural practices and living style. It may be also the reason of variation of the basic reproductive ratio (*R*_0_) values across the selected states, as reported in this study.

As already mentioned, for studying the SEIRD mode, the parameters like-incubation rate, mortality rate, *R*_0_ and recovery rate are required to be calculated. For a given epidemic *R*_0_ is fixed,it denotes the number of secondary infections are generated from a seed infection. The value of *R*_0_ is calculated when an epidemic is in its growing stage or early stage. We have employed a maximum likelihood estimation method depending on daily incidence and generation time outlined by Obadia *et al*. and Zhang *et al*.[3, 4]. The value of *R*_0_ is estimated to be 1.917, which is within the ranges reported in various studies [4, 5, 6, 7, 8, 9, 10]. The range is expected to become narrower if analysed with more data. Containment measures may change the effective reproductive ratio *R*_*e*_ and in India it is currently between 1.43-1.55 as estimated in this study, for incubation periods of 5 days and serial intervals of 8 and 10 days respectively. The incubation period (*d*_*i*_) for COVID-19 has been estimated between 2-14 days (WHO). In many studies it is suggested that mean incubation period is between 5-8 days [11, 12, 6, 13, 14]. Also the average incubation period is estimated to be 5.2 in a Wuhan based study by Li *et al*. [15] as notified in Worldometer. We have taken a range of 5-7 days, for mean incubation period motivated by these studies and Indian studies [16, 17]. The other important parameter for modelling the epidemic is to find the period of infectiousness. For India, without having relevant data, we have resorted to look upon other studies. The other studies suggest mean infectiousness period (*d*_*if*_) is between 3 − 5 days [7, 18]. It is to also remember that *d*_*if*_ for India will be on lower side, i.e., will be close to 3 days as lock-down, social distancing, contact tracing and isolation of the infected or exposed people are done from very early stage of the disease. However, for a comparative study we have considered the values 3, 4, 5 days for analysing the model. The mortality rate of COVID-19 is generally low. In India it is only 3.1% of the total infected population has deceased as of current data. We have calculated the mortality rates for 8 states and for India using total number of deaths and total confirmed cases over a period.

This study shows, India has done much better job than some other countries where the number of infections per million has been quite large. The infection in India will decline around 10-12 Spetember. Other states also may see flattening of the curve between June-August. With a Gaussian fit of the real data, India will see a peak around mid August if no drastic change in the disease dynamics appears. The total number of infections would have been at least three times higher than the present scenario with containment measures in place for India.

Finally, a comparison of progress of COVID-19 with two countries with higher and lower infections than India are done. Russia has more infections than India at the moment and Italy has already moved to almost zero incidence limit. The comparison shows that due to huge population, India’s total number of infections may turn out to be higher than these countries but the rate of infections per million will be quite low. This would mean that the number of cases with severe illness will be under control, which not going to pose a serious challenge to the Government based health care providers. However, there will still be challenges in states like Maharashtra and Delhi due to high rate of infections.

## 2. Model

We have employed a standard epidemiological model for infectious disease, namely SEIRD model.

Let us briefly discuss the model.

### 2.1. SEIRD Model

To model and analyze the data the Susceptible(S) - Exposed(E) - Infectious(I) - Recovered(R) - Dead(D) model is used. Since COVID-19 has a latent phase during which the individual is infected but not yet infectious. This delay between the acquisition of infection and the infectious state can be incorporated within the simpler SIR model by adding a latent/exposed population, E, and letting infected (but not yet infectious) individuals move from S to E and from E to I. *D* is the dead population.

The dynamical equations for SEIRD model are :

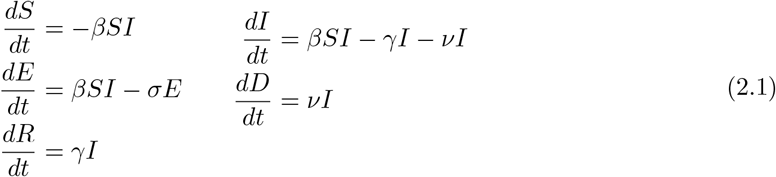

For simulation purpose we cast the above equations in the following discrete time progression form

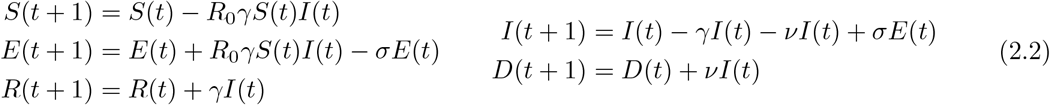

with initial conditions *S*(0) > 0, *E*(0) ≥ 0, *I*(0) ≥ 0 and *R*(0) ≥ 0. Constants *β, γ, σ, ν* are defined in Section 3. Mortality rate can be calculated from Covid-India data and is related to *ν*. The recovery rate is inverse of mean infectiousness period, that is average time to get cured from the onset of symptoms. The recovery rate is taken as 3, 4 and 5 days, as suggested by Bi *et al*. [6] from the studies based on Wuhan, inverse of which gives *γ* in equation (2.1).

### 2.2. Data source

The number of people infected, recovered and deceased daily from 14th March to 12th June from Indian government’s website [19, 20] that are publicly available and data from public domain [21, 22] was obtained. From this available data, few pre-processing were carried out for the SEIRD based model calculations. The population density was collected from 2011 census [23].

## 3. Methodology

Let us now describe the definitions and method of computation of the parameters for studying the population dynamics with the help of SEIRD model.

### Basic Reproductive Ratio. (R_0_)

The reproduction rate *R*_0_ can be calculated with the daily cases starting from day 1 (March-14). Assuming that daily incidence obeys an approximate Poisson distribution, one employs Maximum Likelihood Estimate method. Given observation of (*N*_0_, *N*_1_, …, *N*) incident cases over consecutive time units, and a generation time distribution *w, R*_0_ is estimated by maximizing the log-likelihood

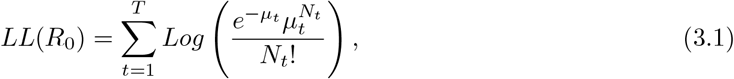

Where 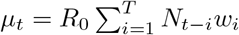. The random vector *w*_*i*_, can be estimated as mean doubling time. *w* is also called serial interval distribution [11, 24].

*R*_0_ calculated for data upto 23 March produced a value between 1.5 − 4 [10]. *R*_0_ produced for data up to 24 May, is 2.093 and *R*_0_ = 1.917 for data up to 12 June.

### Serial Interval. (T_g_)

The serial interval use to be the sum of incubation period and infectious period. Incubation period is the number of days after which a person develops symptoms after being exposed to an infected. Whereas infectious period is defined as the number of days an infected person remains infectious or can infect another person, after getting infected. Doubling time is defined as time period in which the cumulative infection doubles [25, 26]. Doubling Time is a function of both *T*_*g*_ and *R*_0_ as 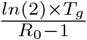 [27].

### Effective Reproductive Ratio. (R_e_)

This is a dynamical quantity that reflects on average during a pandemic the number of person gets infected by an infected person. *R*_*e*_ takes care the effect of measures taken to prevent the spread of the infectious disease. Therefore it will vary as time progresses and lockdown, social distancing, contact tracing etc. are being employed by Government and local administrations. The effective reproductive ratio can be calculated in several ways [3] using a detailed data that can trace the secondary infections generated by a primary infector. However, in absence of such data we have employed a different method depicted by Lipsitch [28]. The method uses average latent period (incubation period) *d*_*i*_, average serial interval *T*_*g*_, growth rate *λ* of the epidemic when it is in exponential phase. The reproductive number then can be estimated as

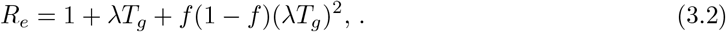

where *λ* = ln [*Y* (*t*)]*/t*, and *f* is the ratio of the mean latent period, i.e., time from infection to onset of infectiousness, to the serial interval or doubling time in our case. *T*_*g*_ is taken to be the sum of the mean latent period and the mean duration of infectiousness.

We have seen the effective reproductive ratio has reduced to 1.55 on the 90th day from 1.84 (on 55th day) currently with *T*_*g*_ = 10 days. For *T*_*g*_ = 8 days, the value of *R*_*e*_ has been calculated equal to 1.43.

### Recovery Rate. (γ)

The recovery rate is defined as the inverse of infectious period *d*_*if*_. The WHO data and other studies [14, 29, 7, 6] indicate the infectious period is between 3-5 days.

### Incubation rate. (σ)

Incubation rate is calculated by taking inverse of mean incubation period *d*_*i*_. WHO data and other studies [30, 11, 13, 29, 7, 6] indicate the incubation period is between 5-7 days.

### Mortality Rate. (ν)

The mortality rate is calculated as number of individuals died on a given period of time divided by the total infected in that period [17].

## 4. Results

We have chosen 8 states based on their population density and COVID-19 infected per million population, as shown in Figure (1). We have omitted low population - high infection, as it does not exist in Indian scenario. We have considered population density > 500*/km*^2^ as high and < 300*/km*^2^ as low. On the other hand, number of infection > 500*/*10^6^ people, is treated as high and < 100*/*10^6^ is considered as low in our distribution.

**Figure 1:**
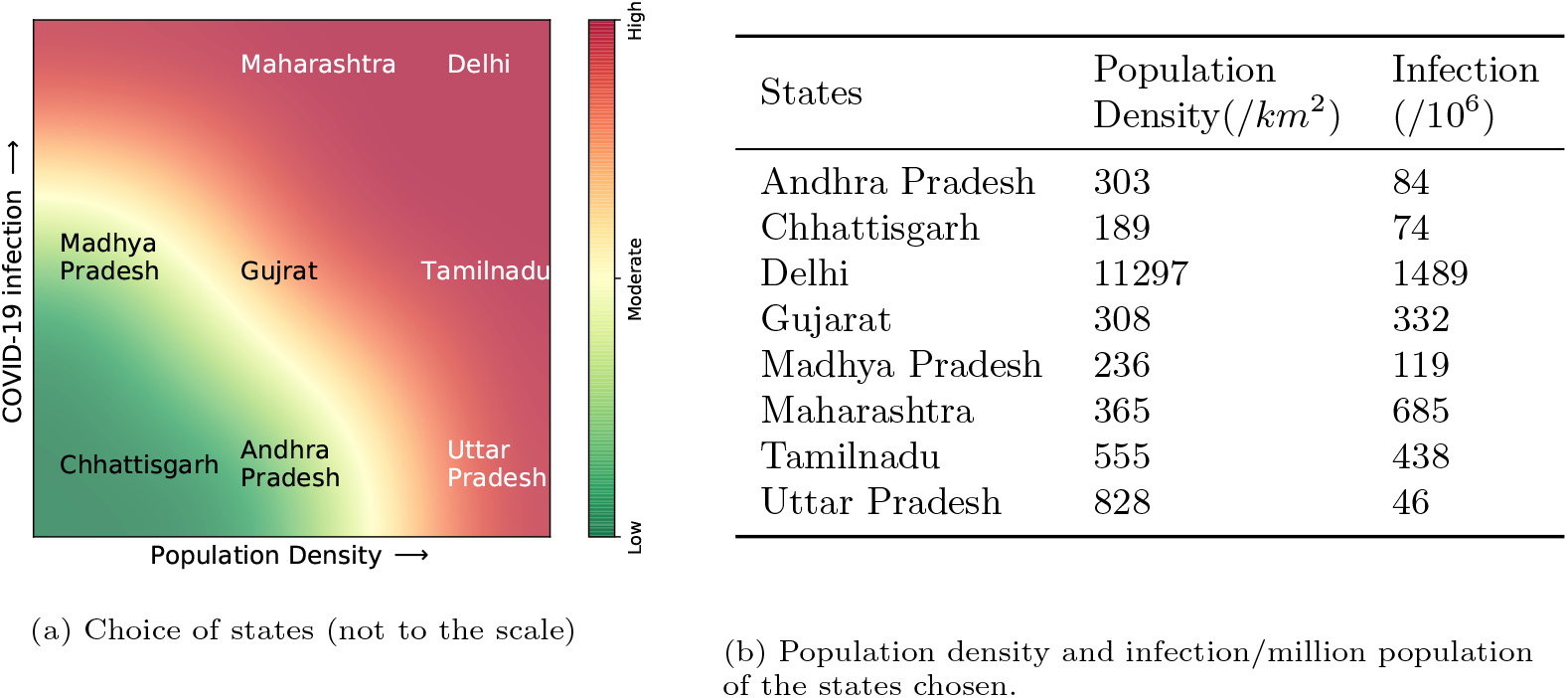
Choice of states.

Previous results suggest average incubation period of SARS-CoV-2 virus between 5 to 7 days [11, 31, 6]. So, we have studied the scenario for incubation period of 5(best case scenario), 6 and 7(worst case scenario) days for each state. Also for each incubation period, we have calculated the infection for *γ*= 3,4 & 5. Previous results suggest incubation period of SARS-CoV-2 virus between 5 to 7 days [11, 31, 6]. So, we have studied the scenario for incubation period of 5(best case scenario), 6 and 7(worst case scenario) days for each state. Also for each incubation period, we have calculated the infection for *γ*=3,4 & 5.

### 4.1. Scenario in selected states

*Andhra Pradesh (AP)*,. a state in Deccan peninsula of India, has moderate population density and low covid infection, as shown in table (1b). The SEIRD results for *σ* = 5, 6 & 7 and *γ* = 3, 4, & 5 is shown in Figure (2). Detailed data are shown in Table (1a). According to our calculations, *R*_0_(*India*) is 2.439. Our calculations also shows the pandemic should over in AP by 89 to 134 days. *These days correspond to the best and worst scenarios in terms of temporal duration of the disease*. 6.6% to 11.0% of susceptible population may be *actively* infected in AP.

**Table 1:** Result for SEIRD Model

| $R_0=2.439$ | | | $R_0=2.414$ | | | $R_0=2.503$ | | |
| --- | --- | --- | --- | --- | --- | --- | --- | --- |
| $\sigma(1/\text{days})$ | $\gamma(1/\text{days})$ | $I_{max}(\text{days})$ | $\sigma(1/\text{days})$ | $\gamma(1/\text{days})$ | $I_{max}(\text{days})$ | $\sigma(1/\text{days})$ | $\gamma(1/\text{days})$ | $I_{max}(\text{days})$ |
| 5 | 3 | 89 | 5 | 3 | 85 | 5 | 3 | 101 |
|  | 4 | 101 |  | 4 | 97 |  | 4 | 115 |
|  | 5 | 113 |  | 5 | 108 |  | 5 | 129 |
| 6 | 3 | 99 | 6 | 3 | 94 | 6 | 3 | 113 |
|  | 4 | 111 |  | 4 | 107 |  | 4 | 128 |
|  | 5 | 123 |  | 5 | 118 |  | 5 | 141 |
| 7 | 3 | 108 | 7 | 3 | 104 | 7 | 3 | 124 |
|  | 4 | 121 |  | 4 | 116 |  | 4 | 139 |
|  | 5 | 133 |  | 5 | 128 |  | 5 | 153 |
| (a) Andhra Pradesh |  |  | (b) Chhattisgarh |  |  | (c) Delhi |  |  |
| $R_0=2.339$ | | | $R_0=2.505$ | | | $R_0=2.087$ | | |
| $\sigma(1/\text{days})$ | $\gamma(1/\text{days})$ | $I_{max}(\text{days})$ | $\sigma(1/\text{days})$ | $\gamma(1/\text{days})$ | $I_{max}(\text{days})$ | $\sigma(1/\text{days})$ | $\gamma(1/\text{days})$ | $I_{max}(\text{days})$ |
| 5 | 3 | 95 | 5 | 3 | 99 | 5 | 3 | 116 |
|  | 4 | 109 |  | 4 | 112 |  | 4 | 132 |
|  | 5 | 121 |  | 5 | 125 |  | 5 | 148 |
| 6 | 3 | 106 | 6 | 3 | 110 | 6 | 3 | 130 |
|  | 4 | 120 |  | 4 | 124 |  | 4 | 146 |
|  | 5 | 133 |  | 5 | 137 |  | 5 | 162 |
| 7 | 3 | 117 | 7 | 3 | 121 | 7 | 3 | 143 |
|  | 4 | 131 |  | 4 | 135 |  | 4 | 160 |
|  | 5 | 144 |  | 5 | 149 |  | 5 | 176 |
| (d) Gujarat |  |  | (e) Madhya Pradesh |  |  | (f) Maharashtra |  |  |
| $R_0=2.356$ | | | $R_0=2.416$ | | | $R_0=1.917$ | | |
| $\sigma(1/\text{days})$ | $\gamma(1/\text{days})$ | $I_{max}(\text{days})$ | $\sigma(1/\text{days})$ | $\gamma(1/\text{days})$ | $I_{max}(\text{days})$ | $\sigma(1/\text{days})$ | $\gamma(1/\text{days})$ | $I_{max}(\text{days})$ |
| 5 | 3 | 95 | 5 | 3 | 99 | 5 | 3 | 186 |
|  | 4 | 108 |  | 4 | 113 |  | 4 | 212 |
|  | 5 | 120 |  | 5 | 126 |  | 5 | 236 |
| 6 | 3 | 105 | 6 | 3 | 111 | 6 | 3 | 208 |
|  | 4 | 119 |  | 4 | 125 |  | 4 | 235 |
|  | 5 | 132 |  | 5 | 139 |  | 5 | 260 |
| 7 | 3 | 116 | 7 | 3 | 122 | 7 | 3 | 230 |
|  | 4 | 130 |  | 4 | 136 |  | 4 | 257 |
|  | 5 | 143 |  | 5 | 150 |  | 5 | 284 |
| (g) Tamilnadu |  |  | (h) Uttar Pradesh |  |  | (i) India |  |  |

**Figure 2:**
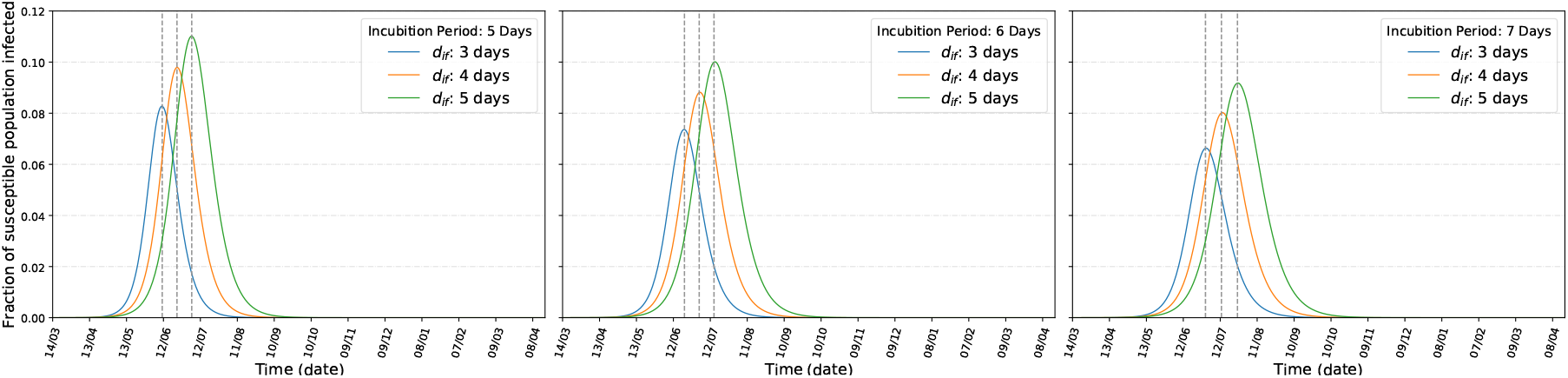
Andhra Pradesh: Infection with incubation period of (left) 5 days, (center) 6 days, (right) 7 days

*Chhattisgarh (CH)*,. a state in central India, has low population density and low covid infection, as shown in table (1b). The SEIRD results are shown in Figure (3). Detailed data are shown in Table (1b). According to our calculations, *R*_0_(*CH*) is 2.414. Our calculations also shows the pandemic should over in CH by 85 to 128 days. 6.5% to 10.8% of susceptible population may be *actively* infected in CH.

**Figure 3:**
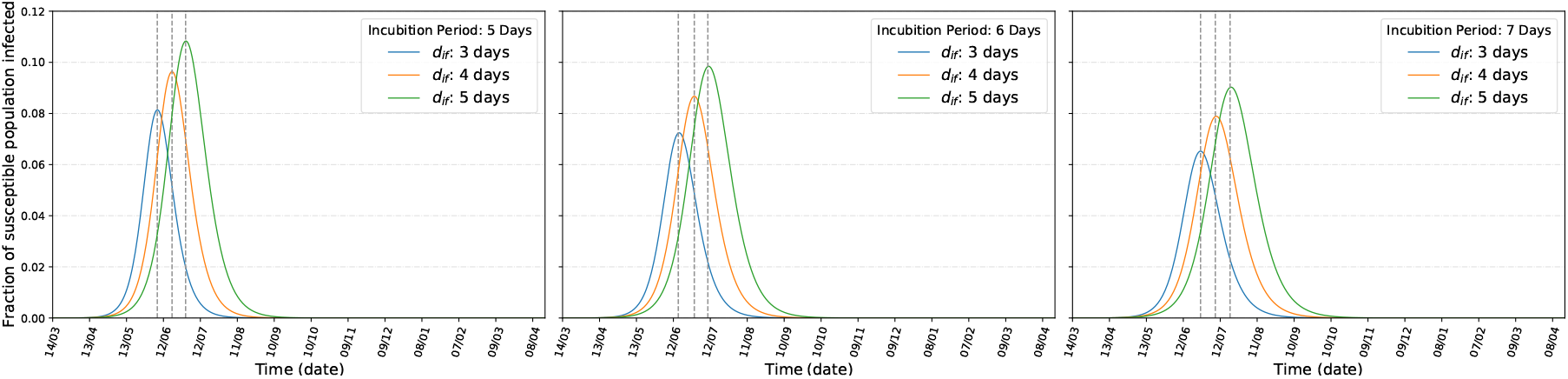
Chhattisgarh: Infection with incubation period of (left) 5 days, (center) 6 days, (right) 7 days

*Delhi*,. the capital of India, is with highest COVID-19 infection and also the most densely populated state of India as shown in Figure (1). SEIRD results for Delhi is shown in Figure (4) and the detailed data is shown in Table (1c). According to our calculation, *R*_0_(*Dl*) is 2.053. Our calculations also shows the pandemic should over in Delhi by 101 to 153 days. 4.8% to 7.99% of susceptible population may be *actively* infected in Delhi.

**Figure 4:**
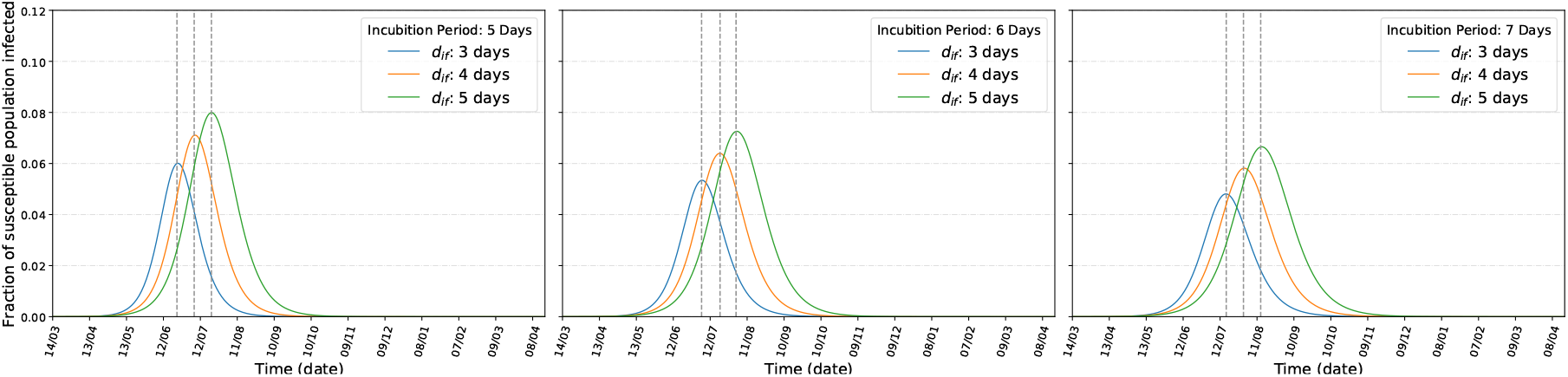
Delhi: Infection with incubation period of (left) 5 days, (center) 6 days, (right) 7 days

*Gujarat(GJ)*,. the western state in India, has moderate population density and also moderate COVID-19 infection, as shown in Figure (1). SEIRD results for Gujrat is shown in Figure (5) and the detailed data is shown in Table (1d). According to our calculation, *R*_0_(*GJ*) is 2.339. Our calculations also shows the pandemic should over in Gujarat by 95 to 144 days. 6.1% to 10.1% of susceptible population may be *actively* infected in GJ.

**Figure 5:**
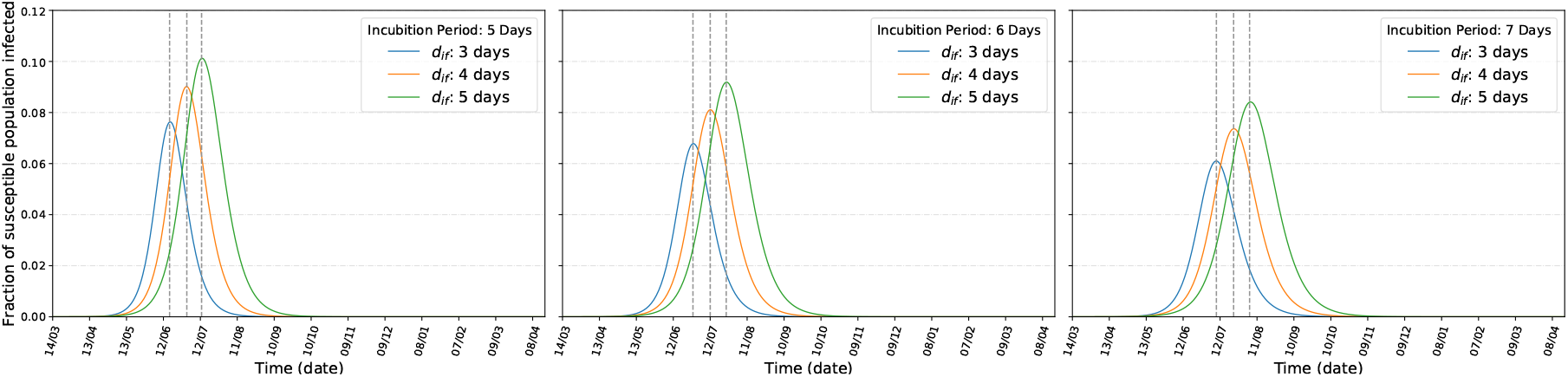
Gujarat: Infection with incubation period of (left) 5 days, (center) 6 days, (right) 7 days

*Madhya Pradesh(MP)*,. the central state in India, has low population density and also moderate COVID-19 infection, as shown in Figure (1). SEIRD results for Madhya Pradesh is shown in Figure (6) and the detailed data is shown in Table (1e). According to our calculation, *R*_0_(*MP*) is 2.303. Our estimation also shows the pandemic should over in MP by 99 to 149 days. 5.9 to 9.9% of susceptible population may be *actively* infected in MP.

**Figure 6:**
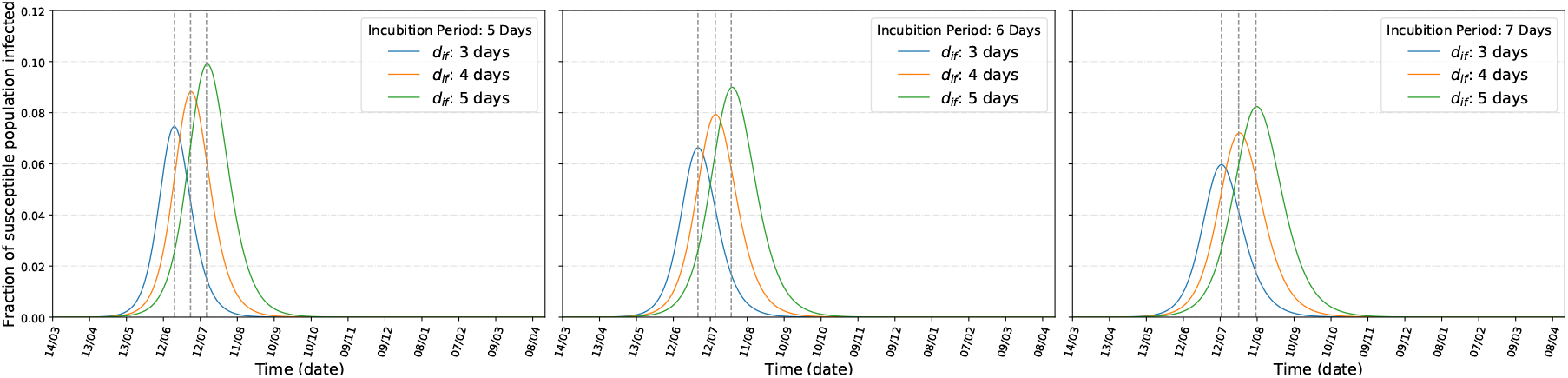
Madhya Pradesh: Infection with incubation period of (left) 5 days, (center) 6 days, (right) 7 days

*Maharashtra(MH)*,. another western state in India, has moderate population density and high COVID-19 infection, as shown in Figure (1). SEIRD results for Maharashtra is shown in Figure (7) and the detailed data is shown in Table (1f). According to our calculation, *R*_0_(*MH*) is 2.087. Our calculation also shows the pandemic should be over in MH by 116 to 176 days. 4.9% to 8.2% of susceptible population may be *actively* infected in MH.

**Figure 7:**
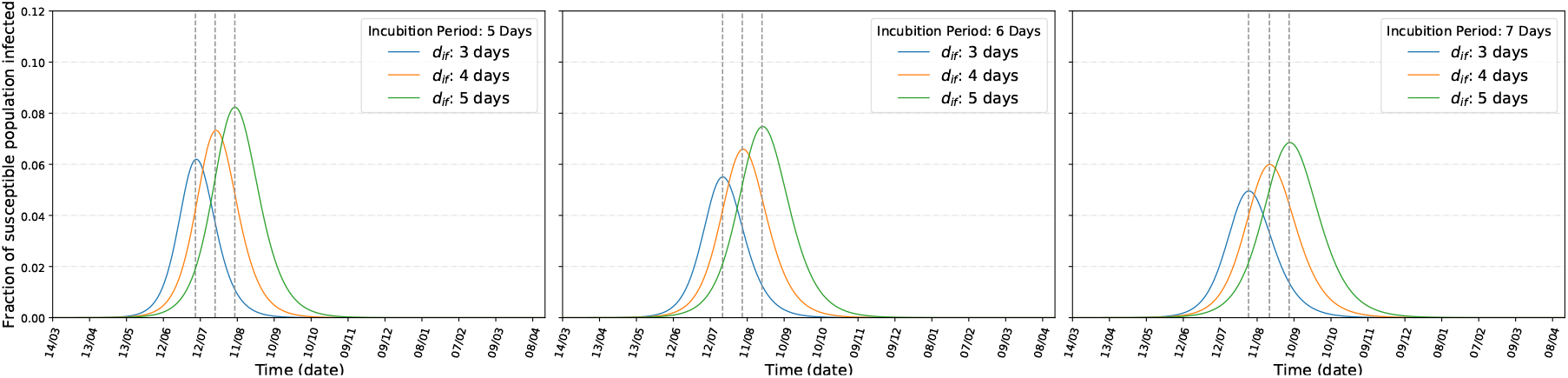
Maharashtra: Infection with incubation period of (left) 5 days, (center) 6 days, (right) 7 days

*Tamilnadu(TN)*,. a southern state in India, has high population density and moderate COVID-19 infection, as shown in Figure (1). SEIRD results for Tamilnadu is shown in Figure (8) and the detailed data is shown in Table (1g). According to our calculation, *R*_0_(*TN*) is 2.356. Our computation also shows the pandemic should over in TN by 95 to 143 days. 6.2% to 10.4% of susceptible population may be *actively* infected in TN.

**Figure 8:**
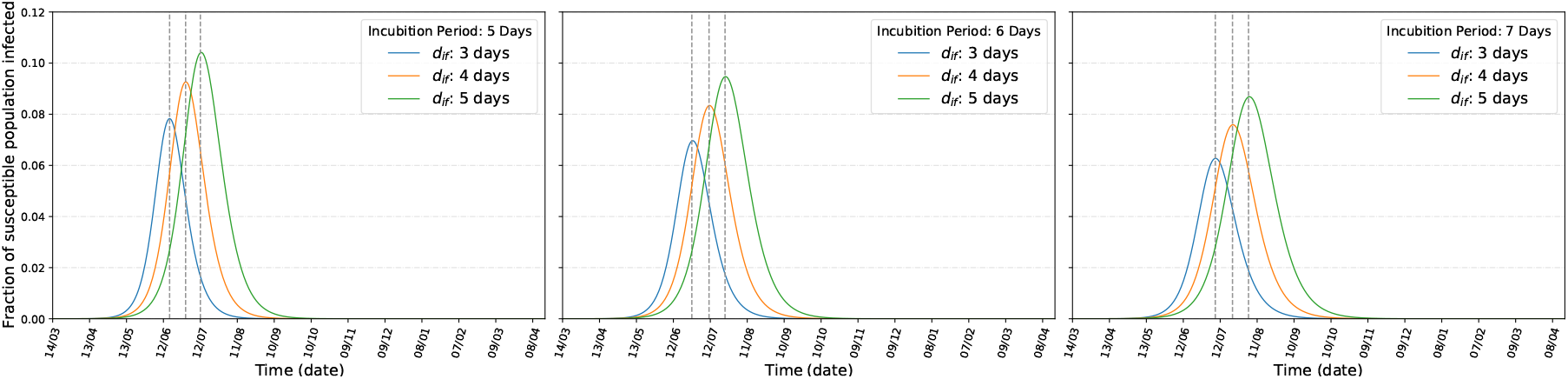
Tamilnadu: Infection with incubation period of (left) 5 days, (center) 6 days, (right) 7 days

*Uttar Pradesh(UP)*,. with highest population in India, has high population density but low COVID-19 infection, as shown in Figure (1). SEIRD results for UP is shown in Figure (9) and the detailed data is shown in Table (1h). According to our calculation, *R*_0_(*UP*) is 2.416. Our study also shows the pandemic should over in MP by 99 to 150 days. 6.5 to 10.8% of susceptible population may be *actively* infected in UP.

**Figure 9:**
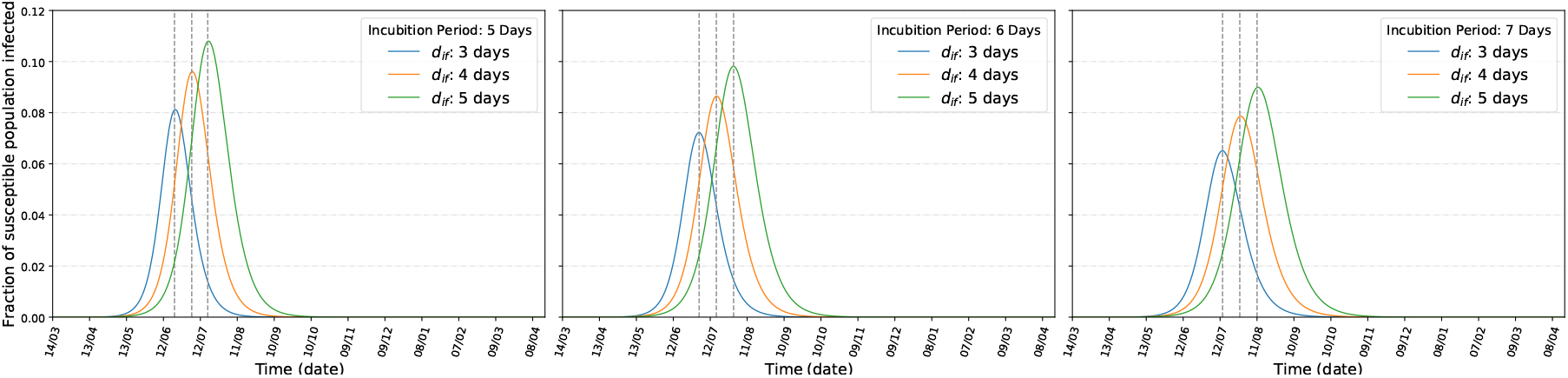
Uttar Pradesh: Infection with incubation period of (left) 5 days, (center) 6 days, (right) 7 days

Finally, India as a whole, has high population density but low COVID-19 infection, with only 30 infected per million population. But, in absolute figure, with 1.3 billion population, this figure is alarmingly high and beyond the capacity of existing medical infrastructure. SEIRD results for India is shown in Figure (10) and the detailed data is shown in Table (1i). According to our calculation, *R*_0_(*India*) is 1.917. Our calculations also shows the pandemic should over in India by 186 to 284 days (depending on incubation and infectiousness periods). 4% to 6.8% of susceptible population may be active at the time of peak appears in India. To get a real estimate of total infected population when the disease will cross the *R*_*e*_ *≤* 1 mark we have calculated *R*_*e*_ for regular periods between 44-90 days, starting from 14th May using equation (3.2). A linear fit shows *R*_*e*_ should go below 1.0 by first week of August (with *d*_*i*_ = 5 and *T*_*g*_ = 10 days) as shown in Figure (11b). This result matches with a Gaussian fit of daily infection, which also reaches its peak around the same time (with approx. a week’s delay), as in Figure (11c). The *R*_*e*_ fit for *T*_*g*_ = 8 with same incubation period shows a peak at 20 days earlier than the peak at Gaussian fit. For the fitting of Figure (11c), we have got correlation functions(*C*) as *C*(amplitude, center)= 0.999, *C*(center, sigma)= 0.997, *C*(amplitude, sigma) = 0.992. These two trends support the plot with 5 days of incubation period and 3 days of infectiousness period for India (10) using the SEIRD model.

**Figure 10:**
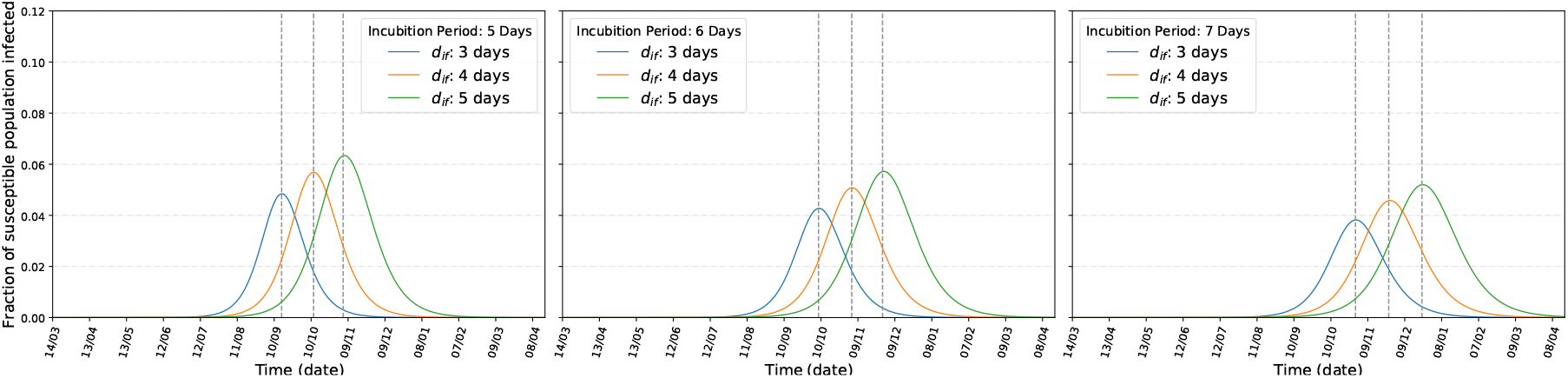
India: Infection with incubation period of (left) 5 days, (center) 6 days, (right) 7 days

**Figure 11:**
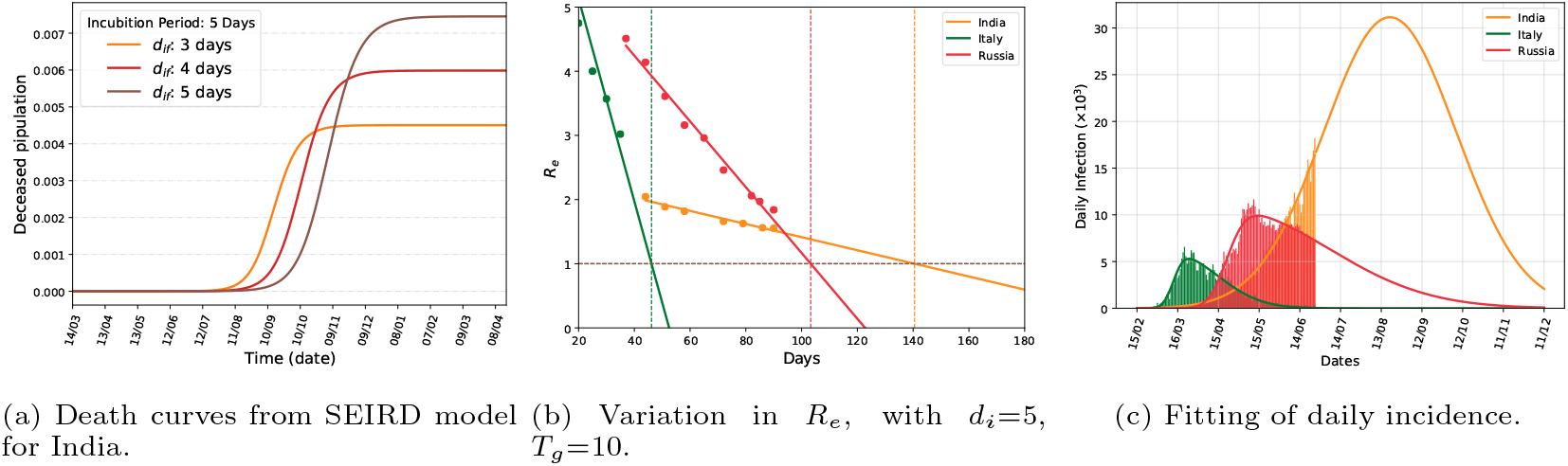
Indian scenario: Comparison with Italy and Russia.

### 4.2. Infection scenario in India

From the Gaussian fit (11c), the total active cases is calculated to be around 5, 12, 700 at the time of peak. Total number of infected people will be roughly 3.1 million at the end of transmission. With the SEIRD model, we can see the peak appears around September middle. From the ICMR data [32], it is observed that the current rate of positive cases is 6.4% of the number of tests done. If the test rate remains the same with a steady increase of 15000 tests per fortnight, the total number of tests in India will reach approximately 25.8 million around mid September. The present rate yields a total 1650560 infected cases in India around mid September. These projections are quite remarkably consistent with the predicted values from the Gaussian fit. From Gaussian fit, the total infected at the peak appears to be approximately 1.612 million. It must be noted, the numbers projected from the Gaussian fit are excluding the asymptomatic cases that are not being reported officially.

The prediction of actual number of infected people from SEIRD model will be grossly overestimated if one assumes that initially the entire population was susceptible. Therefore one needs to determine the actual number of initially susceptible population *S*(0). This can be achieved here with the help of the Gaussian fit. From the plot (10), if we take the peak value corresponding to *d*_*i*_ = 5 and *d*_*if*_ = 3 (this case is the closest to the peak of the Gaussian fit), and compare with the effective active cases predicted from the Gaussian fit, then we get *S*(0) ≈ 10463265. So, initial susceptible population for India is close to only 1% of the total population.

#### Number of deaths

The SEIRD model predicts the peak of the Death curve, with *d*_*i*_ = 5, *d*_*if*_ = 3 will be only 0.45% of the actual susceptible population, see (11a). This projects ≈ 47085 deaths at the peak. This is quite expected as we are having total 16 million infected cases at the peak and with close to 3 percent death rate, the number turns out to be 48000, which is quite close to the value of the same predicted from the model. The total deaths at the end of the pandemic is expected to be around 90000 − 100000.

Finally, we compare our result with others countries. For this calculation, we have taken data from Worldometers [22] from 15th February to 25th June. We have chosen Italy (3,969 cases/million population), where the pandemic is virtually over, and Russia (4,254 cases/million population), where pandemic is still in severe but in decline. As per our discussion in last paragraph, the figure (11c) shows though India has quite low infection rate (364/million population), per day. Also at the peak India will see almost 30000 case which is lower than the countries like USA, Brazil where at the peak the number exceeded well above 30k. The plots of *R*_*e*_ for these three countries are shown in the (3.2). As in both Italy and Russia, the peak of infection has already appeared, we can verify our analysis by comparing the plot with actual dates of peaks. The plots show that Italy has crossed the peak around fourth week of March and Russia has crossed it around third week of May. Strikingly, these are fairly accurate dates what we have observed in reality. For Italy, the peak appeared only ≈ 5 days late, and for Russia the peak appeared approximately 10-14 days earlier. Therefore, there is a high chance that India’s COVID-19 infection peak will also follow the *R*_*e*_ curve plotted above.

## 5. Discussions

This study can be divided into two parts. The first part contains estimation of a few parameters that were used to predict the course of the disease in India and other states from the data. The second part contains the implementation of SEIRD model with the help of the estimated parameters. In particular, using COVID-19 data we have computed the *R*_0_s for India and 8 other states with varying population density and infection status. We have implemented a simple but effective model to explain the growth and decline on the epidemic from Indian context. Our study shows that, for states having low (Chhattisgarh) or moderate (Uttar Pradesh) infection rate the pandemic peaks as early as June, where in for a state like Maharashtra, it may take a month longer.

In view of unavailability of detailed and structured data to prominently project the incubation rate, recovery rate, etc., we have considered a range of values motivated from the trend of the disease in those 8 states and in India. The general trend is, for higher incubation and infectious period the peak appears late.

We have shown, an incubation period of 5 days and an infectious period of 3 days are the most favourable values for which the infection will be under control within 186 days from the onset (assumed to be 14 March in our study) in India. This means the flattening of curve will start around middle of September. If the containment measures are not obeyed and community transmission happens then the effective reproduction ratio will increase and there will be delay in attaining the control over the disease. In the worst scenario the peak may appear around mid December.

A Gaussian plot with the available data for India indicates the flattening of the number of daily incidence will start around mid of August. Although this is earlier by almost three-four weeks than the prediction from SEIRD model but we can get a reasonable idea of expected number of infected at peak. This is calculated to be around 1.6 million. With an early estimation of *R*_0_ as high as 4 before lockdown, the total infected population has been reduced to at least one-third of the early projected values.

As approximately 6.5% of the infected population are required hospital support, the total number of cases of hospitalisations should be around 200 thousands in the most optimistic situation. This is well within the capacity of the health care systems of India.

The states like Chhattisgarh, Karnataka, Kerala have already attained their peaks or on the verge of attaining. In particular, our analysis suggests, Chhattisgarh has already attained its peak infection. This is also consistent with the official data. For other states the peak will be attained within 1-2 months from now. So overall, the situation is improving. This is also indicated from the gradual reduction of *R*_*e*_ values for India in (11b).

The presentation made here have necessary caveats. The computation method for effective reproduction ratio (*R*_*e*_) can be improved with a more detailed data and using a time-dependent method suggested by Wallinga *et al*. [24]. The apparent mismatch of 3-4 weeks of the Gaussian fit and model scenario may be attributed to the fact that the data was plotted from March 14 in Gaussian case, whereas the first case was detected as early as the last week of January. The incubation period, infectiousness period are needed to be calculated from more detailed data identifying an index case and then studying its subsequent transmission channels. That will make the prediction more accurate. An age-structured analysis of the disease would be useful to see the effect of the disease for different age groups.

## Data Availability

All data used are linked and properly cited.

## Acknowledgements

We acknowledge the infrastructural supports provided by IIIT Allahabad where this work has been done during the lock down period.

